# Risk of infection and hospitalization by Covid-19 in Mexico: a case-control study

**DOI:** 10.1101/2020.05.24.20104414

**Authors:** Jaime Berumen, Max Schmulson, Jesús Alegre-Díaz, Guadalupe Guerrero, Jorge Larriva-Sahd, Gustavo Olaiz, Rosa María Wong-Chew, Carlos Cantú-Brito, Ana Ochoa-Guzmán, Adrián Garcilazo-Ávila, Carlos González-Carballo, Erwin Chiquete

## Abstract

**Objective:** During the onset of a novel epidemic, there are public health priorities that need to be estimated, such as risk factors for infection, hospitalization, and clinical severity to allocate resources and issue health policies. In this work we calculate the risk of infection and hospitalization by Covid-19 conferred by demographic, lifestyle, and co-morbidity factors.

**Material and methods:** This is a case-control study including the tested individuals for SARS-Cov-2 by RT-PCR officially reported by the Health Secretary of Mexico from January 01 to May 8, 2020 (102,875 subjects). Demographic (sex, age, foreign and immigrant status, native speaking, place of residence), life-style (smoking), and co-morbidities [diabetes, obesity, high blood pressure (HBP), asthma, immunosuppression, chronic obstructive pulmonary disease (COPD), cardiovascular disease other than HBP, chronic kidney disease (CKD), and other not specified diseases (other diseases)] variables were included in this study. The risk of infection and hospitalization conferred by each variable was calculated with univariate (ULR) and multivariate (MLR) logistic regression models.

**Results:** The place of residence (OR=4.91 living in Tijuana City), followed by advanced age (OR=6.71 in 61-70 years-old), suffering from diabetes (OR=1.87) or obesity (OR=1.61), being male (OR=1.55), having HBP (OR=1.52), and notoriously being indigenous (OR=1.49) conferred a higher risk of becoming infected by SARS-CoV-2 in Mexico. Unexpectedly, we found that having asthma (OR=0.63), immunosuppression (OR=0.65) or smoking (OR=0.85) are protective factors against infection, while suffering from COPD does not increase the risk for SARS-CoV-2 infection. In contrast, advanced age (OR=11.6 in ≥ 70 years-old) is the main factor for hospitalization due to Covid-19, followed by some co-morbidities, mainly diabetes (OR=3.69) and HBP (OR=2.79), being indigenous (OR=1.89), male sex (OR=1.67) and the place of residence (OR=4.22 for living in Juarez City). Unlike the protective risk against infection, immunosuppression (OR=2.69) and COPD (OR=3.63), contribute to the risk of being hospitalized, while having asthma (OR=0.7) also provides protection against hospitalization.

**Conclusions:** In addition to confirming that older age, diabetes, HBP and obesity are the main risk of infection and hospitalization by Covid-19, we found that being indigenous, immunosuppression, smoking and asthma protect against infection, and the latter also against hospitalization.

## INTRODUCTION

Coronavirus disease-2019 (COVID-19) the name provided to the new severe acute respiratory syndrome-coronavirus-2 (SARS-CoV-2) that broke in the city of Wuhan, Province of Hubei in China, in December 2019, was declared a pandemic by the World Health Organization (WHO) on March 11th, 2020.^1-3^ During the emergence of a novel epidemic, there are public health priorities that need to be estimated such as risk factors for infection, hospitalization and clinical severity to allocate resources and to issue health policies.^4, 5^ In Wuhan-China, the risk of symptomatic infection increased with age at ~4% per year among adults aged 30-60 years;^5^ while in a systematic review, age, sex, previous hospital admissions, comorbidity data, and social determinants of health, were determinants of hospital admission.^4, 6^ In another systematic review and metanalysis of ten papers including 76,993 patients, high blood pressure (HBP), cardiovascular diseases, diabetes mellitus, smoking, chronic obstructive pulmonary disease (COPD), malignancy, and chronic kidney disease (CKD), were among the most prevalent underlying diseases among hospitalized COVID-19 patients.^6^ In a report by the Centers for Disease Control (CDC) among 7,162 cases with COVID-19 in the United States that were analyzed on March 28th, more than a third of patients had one or more underlying health condition, of which the most commonly reported were diabetes mellitus, chronic lung disease, and cardiovascular diseases.^5^ In another metanalysis, older age and a history of smoking were related to progression and deterioration compared to an improvement and stabilization of the COVID-19 illness.^7^ The presence of obesity is also emerging as a risk factor with a six-fold increase risk for severe COVID-19 illness, as well as for mechanical intubation, especially in patients with metabolic associated fatty liver disease.^8^ Obesity appears to be shifting the severity of COVID-19 to younger ages.^9^ Finally, immunosuppression following renal transplantation, and in patients with cancer, is a risk factor for severe disease, although, not necessarily for a worst prognosis.^10^ In fact, adult and children patients with immunosuppression seem to have a favorable disease course, as compared to the general population, suggesting a protective role of a weaker immune response.^11^

COVID-19 hit Mexico on February 27th, 2020 with an in imported case of a subject that traveled to Italy; while the first death was reported on March 18th.^12^ Currently, the 10.2% mortality rate for COVID-19 in Mexico is one of the highest in the world.^12^ However, risk factors for infection and hospitalization, are almost unknown for this country. Therefore, we aimed at investigating such risk factors from a registry of the Health Secretary of Mexico that included the total number of subjects that have been tested with real-time reverse transcriptase-polymerase chain reaction (RT-PCR assay) of nasopharyngeal swabs for SARS-CoV-2.^13^ We have hypothesized that older aged, presence of chronic health issues including obesity, diabetes, asthma, COPD, CKD, HBP and cardiovascular disorders, as well as smoking, are risk factors for infection and hospitalization for COVID-19 in Mexico. In addition, we also wanted to determine if immunosuppression, would be a protective factor as it has been previously suggested, and to explore if the indigenous population in Mexico would be at a similar risk for COVID-19 as the rest of the population.

## MATERIALS AND METHODS

**Design, study population and setting**. This is a case-control study including the tested individuals for SARS-Cov-2 by RT-PCR officially reported by the Health Secretary of Mexico from January 01 to May 8, 2020 (102,875 subjects).^13^ This registry collects demographic and clinical information from the 475 Respiratory Disease Monitoring Units (USMER) of the Viral Respiratory Disease Epidemiological Surveillance System, located in health centers or hospitals throughout the country and includes all national health systems (IMSS, ISSSTE, SS, SEDENA, SEMAR, others). The information contained in that database corresponds only to the data obtained from the epidemiological study of suspected viral respiratory disease at the time the person is interrogated in those medical units. According to the clinical diagnosis of admission, it was considered whether the patient could be managed ambulatorily or had to be hospitalized. However, the database does not include clinical symptomatology or evolution during the stay in the medical units.

No informed consent was obtained as this was not an interventional study nor a direct survey of the study subjects. This study was based solely on the analysis of a public national registry of subjects that have been tested for SARS-CoV-2 RT-PCR in Mexico. In addition, no personal identifications are present in this dataset therefore there is no bridge in the privacy of the study subjects. However, the Research and Ethics Committee of the Faculty of Medicine of the Universidad Nacional Autónoma de México was consulted, and they replied that ethical approval was not required.

**Main outcome measures**. Demographic (sex, age, foreign and immigrant status, native speaking, place of residence), life-style (smoking), and co-morbidities [diabetes, obesity, HBP, asthma, immunosuppression, COPD, cardiovascular disease other than HBP, CKD, and other not specified diseases (other diseases)] variables were included in this study. Speaking indigenous languages/dialects was used as a surrogate marker of indigenous Mexican population.

For sex, women were considered as the reference group. Age was grouped by age-ranges and patients 20 years-old or younger were considered the reference group. For place of residence, all metropolitan areas with more than 1 million inhabitants were included, and individuals from the rest of the country’s cities were considered the reference group. For all other variables that include a yes or no answer, the “no” response was considered as the reference value. The risk of infection or hospitalization conferred by each variable was calculated with univariate (ULR) and multivariate (MLR) logistic regression models.

### Statistical analysis

Numerical variables were described with medians and interquartile range (IQR) or means and standard deviations (SD). The significance of differences between the groups (cases and controls) was assessed with the Mann-Whitney U-test or the *t*-test. The association of significant variables with infection or hospitalization was explored using ULR and MLR logistic regression models. The association was expressed as the odd ratio (OR) and 95% confidence interval (CI), and the contribution to the variability of be infected or hospitalized was expressed as adjusted r^2^. Variables with p<0.2 in the ULR analysis were considered for entry in MLR models. Confounders were defined as those variables for which the percentage difference of β coefficient between the adjusted and non-adjusted variables in the MLR model were higher than 10% (p>0.1). All statistical tests were two-sided. The statistical analyses were conducted using SPSS version 20 software (SPSS Inc., Chicago, IL, USA).

## RESULTS

### Demographic and clinical characteristics of studied population

A total of 102,875 registered individuals tested for SARS-CoV-2 RT-PCR, were included in the study; 31,522 (30.6%) were positive and 71,353 (69.4) had a negative result. The demographic characteristics of the cases and controls are presented in Table 1. Positivity for SARS-CoV-2 was higher in men than women (35.2% vs. 25.9%, p<0.001), and increases progressively with age, from 10.6% in individuals in the ≤20 year-old group, up to 44.2% in the 61-70 year-old group (p<0.001). Interestingly, the percentage of positivity was higher in native Mexican population than in the rest of individuals (39.5% vs. 30.5%, p<0.001), and much lower in foreigners than in nationals (19.4% vs. 30.8%, p<0.001). On the other hand, positivity in the US-border cities (Tijuana and Juarez) and Cancun, about 60% of the individuals that were explored, was higher than that in Mexico City (38.6%), the State of Mexico (44.5%), and the rest of the country (25.9%). The positivity-rate in individuals with co-morbidities such as diabetes, obesity and hypertension was much higher than the one in subjects without these co-morbidities, mainly with diabetes (43% vs. 28.7%, p<0.001); and it is striking that in asthmatic or individuals with immunosuppression, as well as in smokers, the opposite was found, a lower infection rate compared to those with these factors (see Table 1).

**Table 1.** Demographic and clinical characteristic of studied population (n=102,875)^*^

| Variable | Value | Covid-19 confirmed<br>% (n) |  | Hospitalized for Covid-19<br>% (n) |  |
| --- | --- | --- | --- | --- | --- |
|  |  | No<br>(n= 71,353) | Yes<br>(n= 31,522) | No<br>(n= 18,832) | Yes<br>(n= 12,690) |
| Demographic and Life-style factors |  |  |  |  |  |
| Gender | Woman | 74.1 (37,411) | 25.9 (13,081) | 66.9 (8,748) | 33.1 (4,333) |
|  | Man | 64.8 (33,942) | 35.2 (18,441)<sup>f</sup> | 54.7 (10,084) | 45.3 (8,357)<sup>f</sup> |
| Age | <= 20 years | 89.4 (6,880) | 10.6 (813) | 82.5 (671) | 17.5 (142) |
|  | 21-30 years | 78.5 (14,602) | 21.5 (3,990) | 84.1 (3,355) | 15.9 (635) |
|  | 31-40 years | 72.1 (17,917) | 27.9 (6,923) | 76.8 (5,316) | 23.2 (1,607) |
|  | 41-50 years | 65.5 (14,087) | 34.5 (7,425) | 61.4 (4,556) | 38.6 (2,869) |
|  | 51-60 years | 59.2 (9,089) | 40.8 (6,257) | 47.3 (2,957) | 52.7 (3,300) |
|  | 61-70 years | 55.8 (4,698) | 44.2 (3,727) | 34.5 (1,286) | 65.5 (2,441) |
|  | >70 years | 63.1 (4,080) | 36.9 (2,387)<sup>f</sup> | 28.9 (691) | 71.1 (1,696)<sup>f</sup> |
| Foreign | No | 69.2 (70,354) | 30.8 (31,281) | 59.6 (18,652) | 40.4 (12,629) |
|  | Yes | 80.6 (999) | 19.4 (241)<sup>f</sup> | 74.7 (180) | 25.3 (61)<sup>f</sup> |
| Native speaking | No | 69.5 (70,657) | 30.5 (31,067) | 60 (18,631) | 40 (12,436) |
|  | Yes | 60.5 (696) | 39.5 (455)<sup>f</sup> | 44.2 (201) | 55.8 (254)<sup>f</sup> |
| Immigrant | No | 69.3 (71,188) | 30.7 (31,483) | 59.7 (18,803) | 40.3 (12,680) |
|  | Yes | 80.9 (165) | 19.1 (39)<sup>f</sup> | 74.4 (29) | 25.6 (10)<sup>a</sup> |
| Place of residence | Rest of the country | 74.1 (37,853) | 25.9 (13,206) | 63.1 (8,331) | 36.9 (4,875) |
|  | Mexico City | 61.4 (13,875) | 38.6 (8,705) | 66.5 (5,792) | 33.5 (2,913) |
|  | State of Mexico | 55.5 (6,758) | 44.5 (5,418) | 46.7 (2,531) | 53.3 (2,887) |
|  | Monterrey | 90.9 (5,656) | 9.1 (563) | 83.3 (469) | 16.7 (94) |
|  | Guadalajara | 91.7 (3,859) | 8.3 (348) | 69 (240) | 31 (108) |
|  | Puebla | 65.1 (1,182) | 34.9 (634) | 50.2 (318) | 49.8 (316) |
|  | Tijuana | 36.9 (756) | 63.1 (1,295) | 41.6 (539) | 58.4 (756) |
|  | Juarez City | 42.6 (291) | 57.4 (392) | 28.8 (113) | 71.2 (279) |
|  | Queretaro | 76.7 (605) | 23.3 (184) | 63.6 (117) | 36.4 (67) |
|  | Cancun | 40 (518) | 60 (777)<sup>f</sup> | 49.2 (382) | 50.8 (395)<sup>f</sup> |
| Smoking | No | 69 (64,180) | 31 (28,774) | 60.1 (17,286) | 39.9 (11,488) |
|  | Yes | 72.3 (7,173) | 27.7 (2,748)<sup>f</sup> | 56.3 (1,546) | 43.7 (1,202)<sup>f</sup> |
| Co-morbidities |  |  |  |  |  |
| Diabetes | No | 71.3 (63,566) | 28.7 (25,638) | 65.6 (16,827) | 34.4 (8,811) |
|  | Yes | 57 (7,787) | 43 (5,884)<sup>f</sup> | 34.1 (2,005) | 65.9 (3,879)<sup>f</sup> |
| Obesity | No | 71.1 (61,238) | 28.9 (24,912) | 61.7 (15,371) | 38.3 (9,541) |
|  | Yes | 60.5 (10,115) | 39.5 (6,610)<sup>f</sup> | 52.4 (3,461) | 47.6 (3,149)<sup>f</sup> |
| HBP | No | 71 (60,386) | 29 (24,699) | 65.2 (16,093) | 34.8 (8,606) |
|  | Yes | 61.6 (10,967) | 38.4 (6,823)<sup>f</sup> | 40.1 (2,739) | 59.9 (4,084)<sup>f</sup> |
| Asthma | No | 69 (67,777) | 31 (30,508) | 59.5 (18,145) | 40.5 (12,363) |
|  | Yes | 77.9 (3,576) | 22.1 (1,014)<sup>f</sup> | 67.8 (687) | 32.2 (327)<sup>f</sup> |
| COPD | No | 69.3 (69,550) | 30.7 (30,764) | 60.5 (18,607) | 39.5 (12,157) |
|  | Yes | 70.4 (1,803) | 29.6 (758)<sup>a</sup> | 29.7 (225) | 70.3 (533)<sup>f</sup> |
| Immunosuppression | No | 69.2 (69,435) | 30.8 (30,966) | 60.2 (18,632) | 39.8 (12,334) |
|  | Yes | 77.5 (1,918) | 22.5 (556)<sup>f</sup> | 36 (200) | 64 (356)<sup>f</sup> |
| Cardiovascular diseases | No | 69.3 (69,110) | 30.7 (30,625) | 60.4 (18,488) | 39.6 (12,137) |
|  | Yes | 71.4 (2,243) | 28.6 (897)<sup>c</sup> | 38.4 (344) | 61.6 (553)<sup>f</sup> |
| CKD | No | 69.4 (69,692) | 30.6 (30,733) | 60.6 (18,622) | 39.4 (12,111) |
|  | Yes | 67.8 (1,661) | 32.2 (789)<sup>b</sup> | 26.6 (210) | 73.4 (579)<sup>f</sup> |
| Other diseases | No | 69 (67,671) | 31 (30,334) | 60 (18,205) | 40 (12,129) |
|  | Yes | 75.6 (3,682) | 24.4 (1,188)<sup>f</sup> | 52.8 (627) | 47.2 (561)<sup>f</sup> |

HBP= High blood pressure, COPD= chronic obstructive pulmonary disease, CKD= chronic kidney disease
\*The percentage was calculated by the totals of each line
a. $p>0.05$ , b. $p<0.05$ , c. $p<0.01$ , d. $p<0.001$ , e. $p<0.001$ , f. $p<0.0001$

### Risk of infection with the virus SARS-Cov-2 in Mexico

Interestingly, the place of residence was the factor that conferred the greatest risk to be infected by the SARS-CoV-2. Places with the highest risk were Tijuana (OR=4.91, CI=4.48-5.38, p<0.001), Cancun (OR=4.3, CI=3.84-4.81, p<0.001) and Juarez (OR=3.86, CI=3,31-4,5, p<0.001), followed by the metropolitan area of the State of Mexico (OR=2.3, CI=2.21-2.39, p<0.001) and Mexico City (OR=1.8, CI=1.74-1.86, p<0.001). Notoriously, people living in Monterrey and Guadalajara have a 3.5 to 3.9 times lower risk of becoming infected than those living in the rest of the country (Table 2). The place of residence explains 9% of the variability of the risk of infection. The age was the second most important risk factor for infection (Table 2). The greatest risk starts in the group of 21-30 years-old, relative to the group of 20 years or younger, with an OR=2.31 (CI=2.13-2.51, p<0.001), and then, progressively increases as the age range rises to the age group of 61-70 years-old, which has a 6.71 times higher risk of becoming infected than young people aged ≤20 years-old. Then, in those over 70 years-old, the risk of infection decreases slightly (OR=4.95, CI=4.53-5.41, p<0.001). Despite conferring a high risk, age only contributes 5.8% of the total variability of the risk of infection. Also, men are 1.55 times more at risk of becoming infected than women, although this factor only explains 1.4% of the risk variability. Interestingly, indigenous subjects were 1.49 times more at risk of becoming infected than the rest of the population, however, this factor contributes very little to the variability of the risk of infection (0.1%). On the other hand, foreigners and immigrants in Mexico had a lower risk of becoming infected by the SARS-CoV-2 (Table 2). Individuals with diabetes (OR=1.87, IC=1.81-1.94, p<0.001), obesity (OR=1.61, IC=1.55-1.66, p<0.001) or HBP (OR=1.52, IC=1.47-1.57, p<0.001) were at higher risk of infection than those without it. Among them, diabetes contributed the most to the variability of the risk of infection (1.5%), and the three factors together contribute only 3.3%. It is interesting to note that patients with asthma (OR=0.63, IC=0.59-0.68, p<0.001) or immunosuppression (OR=0.65, IC=0.59-0.72, p<0.001), as well as smokers (OR=0.85, IC=0.82-0.89, p<0.001), had a lower risk of becoming infected by SARS-CoV-2 virus. In addition, COPD did not contribute to the risk of infection, and cardiovascular diseases other than HBP, very marginally decreased it, while CKD increased the risk of infection. In the multivariate analysis, in which variables with a p≤0.2 in the URL models were entered, all variables but three (immigrant, HBP, CKD) remained in the model. Very noticeable, all the variables that together contributed to the risk of infection, only explained 18.4% of the variability of the risk of infection. This suggests that sociocultural variables, not included in the analysis, such as occupation, means of transportation, and compliance with containment measures could be much more important than the factors analyzed in this work to explain the risk of infection.

**Table 2.**
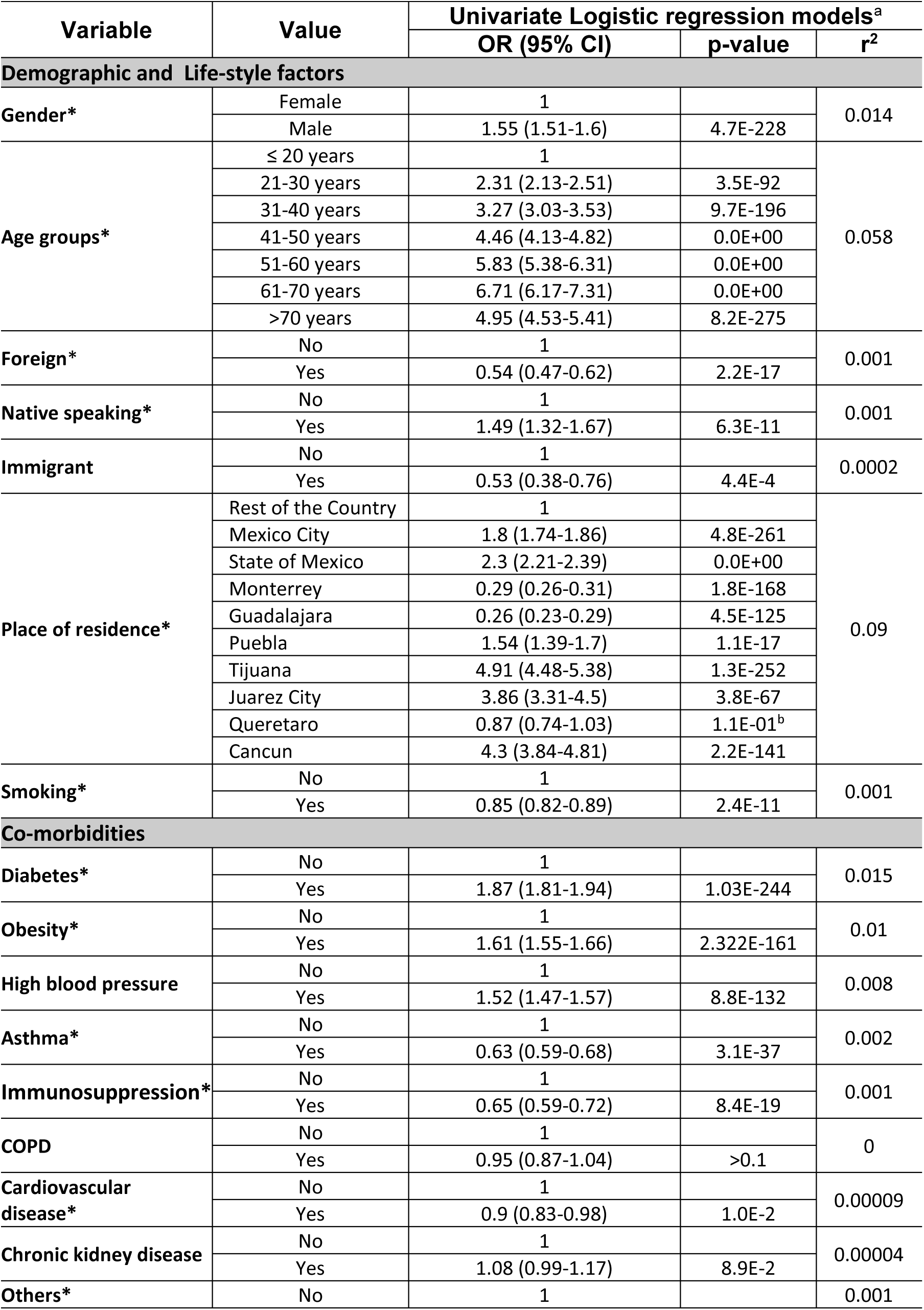
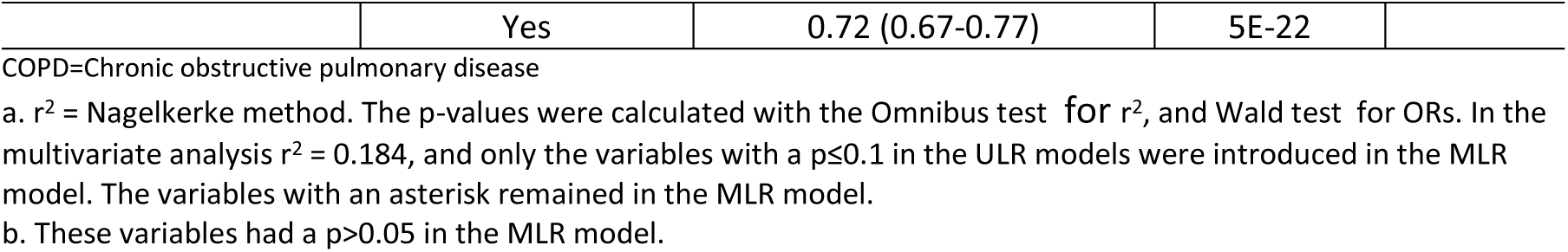
Risk of infection with the virus SARS-Cov-2 in Mexico (n=102,875)

### Risk of hospitalization by Covid-19 in Mexico

For this analysis, the risk of being hospitalized was investigated once the subjects were positive for SARS-Cov-2 RT-PCR (n=31,522). Among them, only 40.3% (n=12,690) were hospitalized. In general, the factors that contributed to the risk of infection also contributed to the risk of hospitalization, with notable exceptions and variations (Table 3). For example, the risk of hospitalization conferred by age, began in the 41-50 years-old group (OR=2.98, CI=2.47-3.59, p<0.001), considering the MLR analysis, and rapidly increased to the >70 years (OR=11.6, CI=9.48-14.19, p<0.001). Unlike the risk of infection, for hospitalization, age contributed much more than the city of residence (18% vs. 4.8%). In fact, age is the main contributing factor for being hospitalized. Living in Juarez confers the highest risk of being hospitalized (OR=4.22, CI=3.38-5.26, p<0.001), followed by the State of Mexico (OR=1.95, CI=1.83-2.08, p<0.001) and Cancun (OR=1.77, CI=1.53-2.04, p<0.001). In contrast, living in Mexico City, Monterrey and Guadalajara, confer lower risk for hospitalization than living in the rest of the country.

**Table 3.**
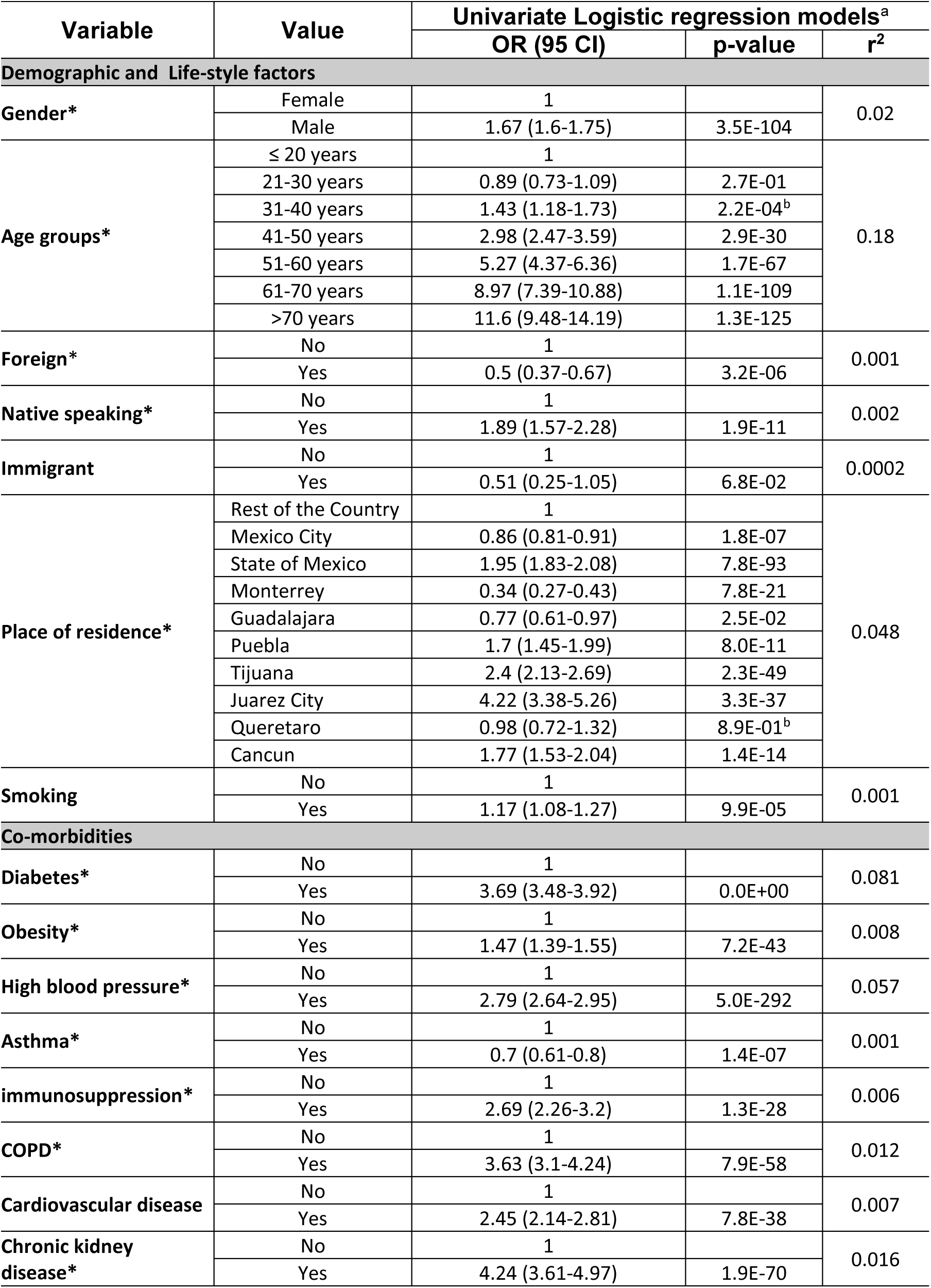
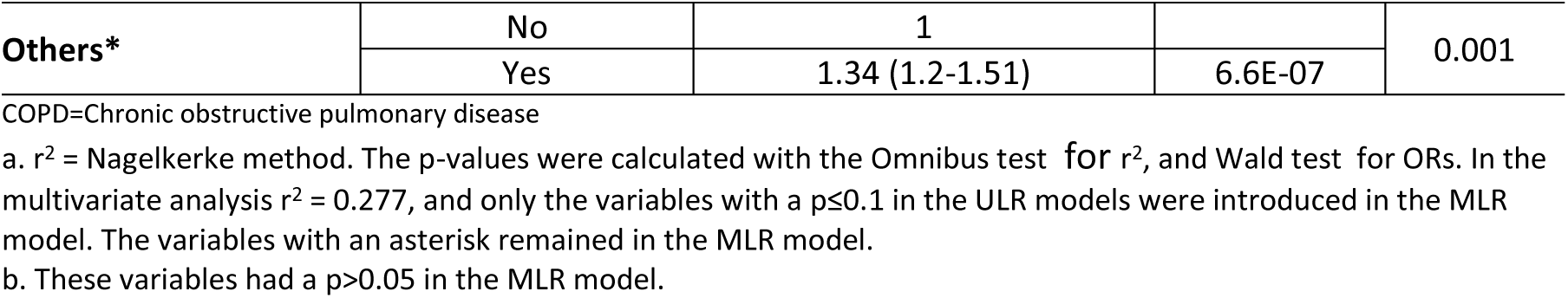
Risk of hospitalization by Covid-19 in Mexico (n=31,522)

Positive individuals for SARS-CoV-2 who speak an indigenous language are almost twice as likely to be hospitalized (OR=1.89, IC=1.57-2.28, p<0.001), suggesting that they consult at a more severe disease stage. Diabetes (OR=3.69, CI=3.48-3.92, p<0.001), contributes more than HBP (OR=2.79, CI=2.64-2.95, p<0.001), and HBP, more than obesity (OR=1.47, CI=1.39-1.55, p<0.001) on the risk of being hospitalized.

On the other hand, factors that protected against infection, like immunosuppression (OR=2.69, CI=2.26-3.2, p<0.001) or other cardiovascular diseases (OR=2.45, CI=2.14-2.81, p<0.001), or that do not contribute to the risk of infection, such as COPD (OR=3.63, CI=3.1-4.24, p<0.001) and CKD (OR=4.24, CI=3.61-4.97, p<0.001), conferred a very high risk for hospitalization. Notoriously, being asthmatic not only protected against infection but also against being hospitalized (OR=0.7, CI=0.61-0.8, p<0.001). In the multivariate analysis all factors remained in the model, except being an immigrant, other cardiovascular diseases, and smoking. The latter conferred a marginal risk for hospitalization in the ULR model. The overall r^2^ value obtained in the MLR model was 0.277, indicating that there are other factors explaining hospitalization, such as the severity of the disease at the time of diagnosis of Covid-19, data that were not recorded in the analyzed registry.

## DISCUSSION

### Key notes

In the current study we have found that place of residence, followed by advanced age, suffering from diabetes, obesity, being male, having BPH, and notoriously being indigenous conferred a higher for infection with SARS-CoV-2 in Mexico. Unexpectedly, we found that having asthma, immunosuppression or smoking are protective factors against infection, while COPD does not increase the risk for SARS-CoV-2. In contrast, advanced age is the main factor for hospitalization for Covid-19, followed by some co-morbidities, mainly diabetes, HBP, being indigenous, male sex and place of residence. Unlike protecting against infection, immunosuppression and COPD, were risk factors for hospitalization, while asthma was a protective factor.

### Strengths and weaknesses

The strengths of this work include the large size of the sample studied, the sampling of individuals at the national level in Mexico, and the analysis of the most important demographic data and comorbidities. However, important weaknesses are the lack of information on the outcomes of the disease and sociocultural data that may be important for the risk of infection, such as occupation, use of mass transit, attendance at mass events, and adherence to containment measures in the country.

### Discussion of findings and contrast with literature

It is interesting to note that the greatest risk of infection and hospitalization is conferred by cities with much trade with the USA (Tijuana and Juarez) and the port of Cancun, which is a massive gateway for foreign tourism to Mexico, both from the USA and Europe. This suggests that at these sites, at the beginning of the pandemic, there was a higher proportion of Covid-19 cases that were imported from abroad and were higher than in other regions of the country, subsequently favoring the pandemic expansion into the region. The metropolitan area, in the center of Mexico, including Mexico City and the State of Mexico, is the second most common place of residence conferring a risk of infection by SARS-Cov-2. However, while people living in Mexico City are less at risk of hospitalization, people living in the State of Mexico have 2-times greater risk than those living in the rest of the country, suggesting that by the time they get the RT-PCR test, the disease is much more advanced.

Because of the age distribution of cases, the Covid-19 pandemic has been characterized as an illness of adults.^14, 15^ Since the initial reports, data on individuals below the age of 20 have been scarce.^16^ Efforts to detect infection in children have been able to identify only a few and most of them present mild symptoms or are asymptomatic and are identified during epidemiological surveillance.^17^ The age disparities in observed cases could be explained by children having lower susceptibility to infection, lower propensity to show clinical symptoms, or both.^18^ This same study estimated that clinical symptoms occur in 25% of infected subjects within the 10-19 age group, rising to 76% in those over 70 years.^18^ In Mexico, the infection distribution according to age is similar to that in China and other countries. Table 1 shows that only 2.6% of all the tests have been performed in people younger than 20, and only 10% were positive. In contrast, close to 40% of individuals older than 60 that were tested, were positive. Furthermore, the risk of becoming infected increased with age, starting from early ages (Table 2). The above data may be related to comorbidities (HBP, diabetes and obesity) that start earlier in life in the Mexican population.^19^ However, this needs to be closely followed because the risk is severe for the age group over 40. On the other hand, the risk of hospitalization follows a well-established dose-response gradient with age, starting at 41 years of age and jumping by decade to almost double for every age group, as shown in Table 3. This pattern has been observed in other countries as well.^20^

According to sex, men are more susceptible to viral infections and worst clinical prognosis,^21^ eliciting lower amounts of interferon alpha (IFN-α) in response to toll-like receptor (TLR) 7 ligands but also higher amounts of the immunosuppressive cytokine interleukin (IL)-10 after stimulation with TLR8 and TLR9 ligands or viruses. More specifically, in a large meta-analysis of Covid-19 a higher incidence in men was reported.^14^ In fact, results from the randomized effects-model meta-analysis revealed a higher male incidence (i.e., 60%) of Covid-19. This proclivity in men to acquire the COVID 19 infection confirm previous accounts for MERS-COV and SARS-COV.^22, 23^ Furthermore, a fatal outcome of Covid-19 infection in men, is consistently found throughout series. Present observations are in line with the notion that males are prone to COVID-19 infection, clinical deterioration and death.^22, 24-26^ Notably, indigenous ethnicity emerged as a risk factor for SARS-CoV-2 infection and the need for hospitalization in this large dataset. However, the fact that indigenous ethnicity is a risk factor for relevant clinical outcomes is a finding that should be investigated in detail.^26^ We have recently described indigenous ethnicity is also a risk factor for COVID-19 death.^25^ It is possible that factors that limit access to healthcare may explain the emergence of this novel risk factor, such as the availability of hospital and ICU beds, dedicated healthcare personnel, medical supplies, as well as geographic distance and time to travel from their communities to hospitals, among other factors. This will be the subject of future analysis.

Globally, patients with SARS-CoV-2 and diabetes or metabolic syndrome had an increased death-rate.^27, 28^ Considering that the burden of diabetes in Mexico is very important increasing the general risk of mortality in at least five fold for every cause,^23^ is a crucial factor that together with obesity might explain the lower age-threshold for the Covid-19 mortality found in this country (>41 years-old); in contrast with other countries reporting increased risk for mortality in those older than 60 or 65.^7^ The process causing damage to kidneys and heart,^29, 30^ might be more lethal in Mexican diabetics. The above may be explained by the frequent presence of uncontrolled levels of glucose and in general, without SARS-CoV-2, they are 30 times more at risk than the rest of the population, to die of kidney failure.^31^ This excess risk causes a larger burden during the pandemic to the already overloaded ICU services and demands for new standards to define the best criteria to manage diabetics in Mexico.^26^

In the current study, obesity was the second most important risk factor for SARS-Cov-2 infection after obesity, however it was not such an important factor for hospitalization. The higher risk for infection is in agreement with findings from China, in which obesity remained a significant risk factor even after adjusting for age, sex, smoking, diabetes, HBP, and dyslipidemia.^8^ However, the fact that in Mexico obesity was less important than other risk factors for hospitalization, is somewhat different than other reports.^22, 32-35^ The connection between obesity and COVID-19 is beyond the scope of the current paper but factors such as attenuation of the immune system and chronic inflammation, have been implicated.^22^ Also, binding of SARS-CoV-2 to the angiotensin converting enzyme 2 receptor (ACE2) occurs at low cytosolic pH values. In the presence of diabetes and conditions such as HBP, obesity, old age, and smoking, cytosolic pH is low, thus the virus may easily enter the cell by attaching to ACE2.^36^ Nevertheless, the higher risk for COVID-19 infection among diabetic and obese subjects in Mexico is an important factor considering that in 2017, this country was number five in population suffering from diabetes in the world, with 12.0 million subjects affected, and has one of the highest obesity prevalence-rates, 32.4% among the 15-74 year-old population-group.^37, 38^ Furthermore, these findings are of great importance for implementing public health strategies for preventing infection with SARS-CoV-2 in this population.^9^

The Centers for Disease Control and Prevention (CDC) guidelines state that patients with moderate to severe asthma could have a greater risk for severe Covid-19 Illness.^27^ In fact asthmatics are at risk for more severe outcomes with other virus infections. It is known that they have a delay innate anti-viral immune responses, with deficiency in lung cell interferon α, β and γ responses related to higher asthma exacerbation severity.^28, 29, 39^ Recommendations for asthmatics during the Covid-19 pandemic are to continue their asthma medication to prevent exacerbations and be lifesaving.^40^ Interestingly, in our study, asthma showed a decreased risk for Covid-19 infection and hospitalization. In Covid-19, the evolution to a critical stage is associated to a cytokine storm, so it could be possible that a regular use of low dosage of corticosteroids or anti-inflammatory drugs, could prevent inflammation and decrease the risk of a critical illness.^41^ In addition, COPD did not increase the risk for Covid-19 infection in Mexico, however, it increased the risk for hospitalization. This is consistent with other studies that reported COPD as a risk factor for a more severe COVID-19 illness.^42, 43^ In addition, the lower risk for infection conferred by immunosuppression herein found, is in contrast with the higher risk for hospitalization of this factor. As mentioned before, clinical deterioration of Covid-19 has been associated to an overactive immune response, and based on this hypothesis, several immunosuppressants are currently studied as potential treatments for severe Covid-19.^44^ Some authors hypothesized that a moderately reduced immune response may play a favorable role in the severity of the diseases due to a lack of significant increase in IL-6.^45^

One of the most intriguing issues is that smoking/nicotine theoretically should be a risk factor for SARS-CoV-2 infection since nicotine affects the ACE2 receptors.^46^ However, our results suggest that smoking is a protective mechanism for Covid-19 infection. It has been published that there is an increased ACE2 expression in the airways of current smokers and those with COPD.^33, 47, 48^ Studies indicate that ACE2 is likely to be the host-receptor for SARS-CoV-2,^49^ but also is involved in the regulation of the renin-angiotensin system (RAS).^50^ Most importantly, ACE2 could have a double mechanism, both protective and pathogenic roles within RAS pathways, and its direct mechanisms in cells, remains unknown.^50^ It has been hypothesized that nicotine, in other health conditions, could influence the cytokine response.^51^ Nicotine is usually confounded with cigarette smoking, since it is the most common mechanism to access the drug. Therefore, public health specialists ought to consider it as a negative risk factor, since smoking is a health hazard.^52^ However, empirical data shows otherwise; nicotine regardless of the pathway utilized to deliver it, may be a protector for the general population,^53^ including for Covid-19 infection. It is clear that promoting smoking in the population would make no sense, however nicotine could be provided as a pharmaceutical product in patches and other forms to protect against Covid-19.

### Conclusions

Based on the public registry of the Secretary of Health of Mexico of all the subject that have been tested with RT-PCR for SARS-CoV-2, we have confirmed that older age, diabetes, HBP and obesity are the main risk of infection and hospitalization for Covid-19. Notably, indigenous ethnicity emerged as a risk factor for SARS-CoV-2 infection and the need for hospitalization in this large dataset. In addition, having immunosuppression, smoking and asthma are protective factors against infection, and the latter one also protects against hospitalization. These findings are important for establishing public health policies and allocate healthcare resources during this pandemic.

## Data Availability

All data generated or analysed during this study are included in this article.

